# User Experience and Therapeutic Alliance in AI-Driven Mental Health Interventions: A Protocol for a Systematic Review of Qualitative Studies

**DOI:** 10.1101/2025.06.22.25330071

**Authors:** Ravi Shankar, Fiona Devi, Xu Qian

**Author notes:** **Corresponding Author:** Dr Ravi Shankar;, Research and Innovation, Medical Affairs, Alexandra Hospital, Singapore Email correspondence.

## Abstract

**Background:** Artificial intelligence (AI) technologies are increasingly being integrated into mental health interventions, but their impact on user experience and the therapeutic alliance remains poorly understood. This protocol outlines a systematic review of methods and applications.

**Objective:** To synthesize qualitative evidence on how AI influences user experience and therapeutic alliance in mental health interventions.

**Methods:** We will search PubMed, Web of Science, Embase, CINAHL, MEDLINE, The Cochrane Library, PsycINFO, and Scopus from inception to June 2025. Qualitative studies exploring user experiences of AI-driven mental health interventions will be included. The ECLIPSE framework will guide the review process. Two reviewers will independently screen studies, extract data, and assess methodological quality using the CASP Qualitative Checklist. Thematic synthesis will be used to analyze and integrate findings across studies. Confidence in the evidence will be assessed using GRADE-CERQual.

**Discussion:** This review will provide insights into the factors shaping user engagement and therapeutic alliance with AI-driven mental health interventions. Findings will inform the design and implementation of AI technologies that optimize user experience and clinical effectiveness. Strengths, limitations, and implications for research and practice will be discussed.

## Introduction

Mental health disorders are a leading cause of disability worldwide, affecting an estimated 970 million people [1]. Despite the high prevalence and burden of mental illness, access to mental health care remains limited due to stigma, cost, and workforce shortages [2]. In recent years, there has been growing interest in leveraging artificial intelligence (AI) to expand the reach and efficiency of mental health interventions.

AI technologies, such as machine learning, natural language processing, and virtual agents, have the potential to transform mental health care delivery. AI-driven interventions can provide personalized, on-demand support at scale, reducing barriers to access and engagement [3]. Emerging applications include chatbots for mental health screening and self-management [4], virtual therapists delivering cognitive-behavioral therapy [5], and smartphone apps for symptom monitoring and prediction [6].

While AI holds promise for improving the accessibility and effectiveness of mental health care, important questions remain about its impact on the user experience. Mental health interventions often rely on a strong therapeutic alliance - the collaborative bond between therapist and client - to promote engagement, adherence, and clinical outcomes [7]. It is unclear how this human-to-human relationship translates to interactions with AI systems, where the “therapist” is a computer algorithm rather than a human clinician.

Early research suggests that users may form emotional bonds with AI chatbots and virtual agents, experiencing them as empathetic, nonjudgmental, and trustworthy [8,9]. However, the depth and stability of these bonds, and their impact on therapeutic processes and outcomes, remains uncertain. Some users report difficulty engaging with AI systems that feel impersonal or lack human qualities like warmth and humor [10]. Concerns have also been raised about the potential for AI to undermine therapeutic alliance by replacing human interaction, dehumanizing care, or eroding trust [11].

Understanding user experiences of AI-driven mental health interventions is critical for designing technologies that optimize engagement, alliance, and clinical benefit. Qualitative research, which explores individuals’ perceptions, attitudes, and behaviors in rich detail, offers valuable insights into the human-computer interaction in mental health care. By synthesizing findings across qualitative studies, we can identify facilitators and barriers to user engagement and therapeutic alliance with AI systems.

To date, no systematic review has comprehensively examined qualitative evidence on user experiences of AI-driven mental health interventions. Prior reviews have focused on quantitative outcomes such as feasibility, acceptability, and effectiveness [12-14], with limited attention to the psychological and relational processes underpinning these metrics. Qualitative evidence has been considered only peripherally, as a supplement to quantitative findings.

This protocol outlines a systematic review designed to address the existing gap by synthesizing qualitative studies focused on user experience and therapeutic alliance in AI-driven mental health interventions. Our aim is to comprehensively understand how users perceive and engage with AI technologies within mental health care, including how these technologies influence the therapeutic process and relationship. Specifically, the review will explore user perceptions and attitudes toward AI-driven mental health interventions, identify factors affecting user engagement and adherence over time, examine how therapeutic alliance is conceptualized and experienced in relation to AI technologies, compare user experiences across different intervention modalities, target problems, and populations, and synthesize recommendations to optimize user engagement and enhance therapeutic alliances within AI-driven mental health interventions.

Findings will provide a framework for understanding the human experience of AI-mediated mental health care, informing the design and implementation of user-centered interventions. Insights into facilitators and barriers to engagement and alliance can guide strategies to build trust, rapport, and collaboration between users and AI systems. Ultimately, this knowledge will help realize the full potential of AI to complement and extend, rather than replace, human-delivered mental health care.

## Methods

This systematic review protocol is reported in accordance with the Preferred Reporting Items for Systematic Review and Meta-Analysis Protocols (PRISMA-P) guidelines [15]. The review has been registered with the International Prospective Register of Systematic Reviews (PROSPERO): CRD42027755998.

### Review Framework

The review will be guided by the ECLIPSE framework [16], which provides a structured approach for examining complex health interventions across multiple dimensions. This includes understanding the Expectations, such as the anticipated outcomes and broader impacts of AI-driven mental health interventions. The Client dimension focuses on the key characteristics of the populations engaging with these interventions. Location refers to the specific settings or contexts in which the interventions are implemented, which may influence their effectiveness and reception. The Impact dimension addresses the experiences, perceptions, and reactions of users and other stakeholders involved. The Professionals component highlights the roles and contributions of those delivering the intervention, including clinicians, developers, or support staff. Lastly, the Service dimension captures the essential elements, delivery strategies, and operational features of the intervention itself. This framework enables a comprehensive exploration of the multifaceted nature of AI-supported mental health care.

This framework aligns with our aim of understanding user experience and therapeutic alliance as embedded within the broader context of AI-driven mental health interventions. By attending to factors at multiple levels - from individual expectations to service design - we can develop a more comprehensive view of how these interventions are perceived and engaged with in real-world settings.

### Eligibility Criteria Inclusion criteria

Studies will be included if they meet the following criteria:

- Qualitative empirical study
- Explores user experiences, perceptions, attitudes towards AI-driven mental health interventions
- AI is a core component of the intervention (e.g., chatbot, virtual agent, machine learning algorithm)
- Mental health intervention targets a specific mental health problem (e.g., depression, anxiety, substance use)
- Includes primary data from users of the intervention (e.g., patients, clients, consumers)
- Uses qualitative methods for data collection (e.g., interviews, focus groups, open-ended surveys) and analysis (e.g., thematic analysis, grounded theory, phenomenology)
- Published in English
- Published in a peer-reviewed journal

### Exclusion criteria

Studies will be excluded if they:

- Do not include an AI-driven intervention (e.g., face-to-face therapy only)
- Do not target a mental health problem (e.g., general wellbeing, physical health)
- Do not report primary qualitative data (e.g., review articles, commentaries)
- Do not focus on user experience (e.g., clinician or developer perspectives only)
- Report only quantitative data (e.g., satisfaction ratings, usage metrics)

Mixed methods studies will be included if they report qualitative findings relevant to the review objectives.

### Search Strategy

We will conduct a comprehensive search across the following electronic databases from inception to June 2025: PubMed, Web of Science, Embase, CINAHL, MEDLINE, The Cochrane Library, PsycINFO, and Scopus. The search strategy will employ a combination of keywords and controlled vocabulary terms (such as MeSH headings) encompassing four key concepts: (1) artificial intelligence, (2) mental health, (3) interventions, and (4) qualitative research. Search terms will be tailored to suit the indexing system of each database. While no date restrictions will be applied, only studies published in English will be included. An example of a search string is as follows:

(“artificial intelligence” OR “machine learning” OR “deep learning” OR “natural language processing” OR chatbot* OR avatar* OR “virtual agent*” OR “conversational agent*”) AND (“mental health” OR “mental illness” OR “mental disorder*” OR depression OR anxiety OR “substance use” OR trauma OR “eating disorder*” OR “serious mental illness”) AND (intervention* OR treatment* OR therap* OR psychotherap* OR “digital health” OR “mobile health” OR mhealth OR ehealth OR telehealth OR telepsychiatry) AND (qualitative OR interview* OR “focus group*” OR ethnograph* OR “grounded theory” OR phenomenolog* OR “thematic analys*” OR “content analys*” OR “framework analys*” OR “discourse analys*” OR “narrative analys*” OR experience* OR perception* OR perspective* OR attitude* OR view*)

We will also hand-search reference lists of included studies and relevant reviews, and conduct forward citation tracking to identify additional eligible studies. Grey literature will be searched via ProQuest Dissertations & Theses, Open Grey, and Google Scholar.

### Study Selection

Two reviewers will independently screen all titles and abstracts identified from the search. Studies appearing to meet inclusion criteria will be obtained in full text and assessed for eligibility by two independent reviewers. Discrepancies will be resolved through discussion or consultation with a third reviewer. Reasons for exclusion at the full-text stage will be documented. The study selection process will be reported using a PRISMA flow diagram [17].

### Data Extraction

Data extraction will be conducted by one reviewer using a standardized form, with a second reviewer verifying all extracted information for accuracy and completeness. Any disagreements will be resolved through discussion and consensus, or by consulting a third reviewer if needed.

The extracted data will include the following categories: (1) study characteristics, such as authors, year, country, aims, setting, and methodology; (2) participant characteristics, including number, age, gender, mental health condition, and experience with the intervention; (3) intervention characteristics, covering the type of AI technology, its intended purpose, delivery format, duration, and intensity; (4) qualitative methods used, including sampling strategies, data collection techniques, and analysis approaches; and (5) key qualitative findings, encompassing user expectations, attitudes, and experiences, therapeutic alliance, factors influencing engagement and adherence, and recommendations for enhancing user engagement and alliance in AI-driven mental health interventions.

Where studies report both quantitative and qualitative methods and findings, we will extract only the qualitative data. We will contact study authors to obtain any relevant data missing from published reports.

### Quality Appraisal

Two reviewers will independently evaluate the methodological quality of all included studies using the Critical Appraisal Skills Program (CASP) Qualitative Research Checklist [18]. This checklist assesses qualitative research across ten domains: a clear statement of aims, appropriate methodology, suitable research design, appropriate recruitment strategy, relevant data collection methods, researcher reflexivity, ethical considerations, rigorous data analysis, clarity of findings, and the overall value of the research. Each study will receive an overall quality rating of high, moderate, or low based on how many of these domains are adequately addressed. While the quality appraisal will inform the overall assessment of the evidence base and help identify potential sources of bias, no studies will be excluded solely based on quality, as even lower-rated studies may offer important insights.

### Data Synthesis

Qualitative findings will be synthesized using a thematic synthesis approach adapted from Thomas and Harden [19], which involves three key stages. First, study findings will undergo line-by-line coding to identify and label meaningful segments of text. Next, these codes will be organized into related categories to develop descriptive themes that capture recurring patterns across studies. Finally, analytical themes will be generated by interpreting and recontextualizing the descriptive themes to address the overarching review questions and provide deeper insights that extend beyond the original findings of the primary studies.

Synthesis will begin with in-depth reading of each study to identify all text (participant quotes and author interpretations) relevant to user experience and therapeutic alliance. Two reviewers will independently code this text line by line, assigning labels that capture its meaning and content. Codes will be organized into related clusters to develop an initial coding framework. This framework will be applied to subsequent studies and refined iteratively as new codes emerge.

In the next stage, codes will be compared and contrasted across studies to develop broader descriptive themes. These themes will summarize patterns and commonalities in how participants perceive and interact with AI-driven interventions. Finally, descriptive themes will be further interpreted and abstracted to generate higher-order analytical themes that directly address the review objectives.

Throughout the process, reviewers will meet regularly to discuss and refine codes, themes, and interpretations. Memos will be used to document decisions and insights. The synthesis will move back and forth between themes and the original studies to ensure findings remain grounded in the data. Negative or disconfirming cases will be sought out to challenge and expand interpretations.

Results will be presented narratively, using text, tables, and figures to summarize themes and relationships. Illustrative participant quotes will be included to support key findings. Analysis will attend to potential differences in user experiences across intervention modalities, target problems, and populations. Recommendations for improving engagement and therapeutic alliance will be synthesized from both participant and researcher suggestions.

### Confidence in the Evidence

Confidence in the review findings will be assessed using the GRADE-CERQual (Grading of Recommendations Assessment, Development and Evaluation–Confidence in the Evidence from Reviews of Qualitative Research) approach [20]. This method evaluates each review finding across four domains: the methodological limitations of the studies contributing to the finding, the coherence of the finding in relation to the underlying data, the adequacy of the data supporting the finding, and the relevance of the data to the specific review question. These assessments will guide the overall confidence rating assigned to each synthesized finding.

Based on these assessments, each finding will be assigned a level of confidence (high, moderate, low, or very low) reflecting the extent to which it is a reasonable representation of the phenomenon of interest.

## Discussion

This systematic review will provide a comprehensive synthesis of qualitative research on user experiences of and therapeutic alliance with AI-driven mental health interventions. By integrating evidence across a range of technologies, populations, and contexts, we aim to develop a nuanced understanding of the human-computer interaction in AI-mediated mental health care.

A key strength of our approach is the use of a robust qualitative methodology to explore the subjective, experiential aspects of engagement with AI interventions. Thematic synthesis will enable us to go beyond a summary of existing findings to generate new theoretical insights and practical recommendations. The ECLIPSE framework ensures we consider the complex, multi-level factors influencing user experience and engagement.

Confidence in the evidence will be carefully evaluated using the systematic and transparent GRADE-CERQual approach. This will help users of the review to interpret the findings and assess their applicability to different contexts. The use of qualitative comparative analysis will highlight potential sources of heterogeneity in user experiences.

Limitations of the review include the restriction to English-language publications, which may underrepresent cultural diversity in user perspectives. The rapid evolution of AI technologies may mean that findings from older studies are less relevant to current applications. Finally, the focus on qualitative evidence precludes any quantitative estimates of engagement, alliance, or effectiveness.

Findings will have important implications for the development and implementation of AI-driven mental health interventions. In particular, insights into facilitators and barriers to engagement and alliance can guide co-design processes to create interventions that are more responsive to user needs and preferences. The review will also identify priorities for future research to address gaps in the evidence base.

From an ethical perspective, the review foregrounds the importance of centering user voices and experiences in the development of AI technologies for mental health. By synthesizing diverse perspectives, we aim to promote a more inclusive and democratic approach to the design and governance of these interventions. At the same time, we acknowledge the potential limitations and risks of AI in mental health care, including issues of privacy, transparency, accountability, and bias.

In conclusion, this systematic review will provide a timely and valuable synthesis of qualitative evidence on user engagement and therapeutic alliance with AI-driven mental health interventions. Findings will contribute to a more nuanced and contextual understanding of the human-computer interaction in mental health care, informing the development of more effective, engaging, and ethical interventions. As AI technologies continue to evolve and penetrate mental health services, this review will help ensure that the user experience remains at the heart of research and practice.

## Data Availability

All data produced in the present work are contained in the manuscript. No additional datasets were generated or analyzed during the current study.

